# Risk factors for invasive Group A Streptococcal infection in children aged 6 months to 5 years, a case-control study, the Netherlands, February-May 2023

**DOI:** 10.1101/2024.11.13.24316742

**Authors:** Elizabeth I Hazelhorst, Catharina E van Ewijk, Cornelia CH Wielders, Margreet JM te Wierik, Susan JM Hahné, Hester E de Melker, Mirjam J Knol, Brechje de Gier

## Abstract

In 2022, an increase in invasive Group A Streptococcal infections (iGAS) was observed in the Netherlands. A particular increase was seen among children, therefore we aimed to assess risk factors for iGAS in children aged 6 months to 5 years. A prospective case-control study was conducted between February and May 2023. We approached parents of notified iGAS cases to complete a questionnaire on exposures during 4 weeks prior to disease onset. Controls were recruited via social media and matched to cases (10:1) on sex and birthyear. Conditional logistic regression was performed to estimate odds ratios (OR) of exposures. For the analysis we included 18 cases and 103 controls. Varicella prior to onset of iGAS disease was reported in 2 (11%) cases and 1 (1%) control (OR: 12.0, 95% CI: 1.1-139.0). Exposure to GAS-like illnesses impetigo, pharyngitis and scarlet fever was reported in 8 (44%) cases and 15 (15%) controls (OR: 7.1, 95% CI: 1.8-29.0). Our findings are in line with previous studies by identifying varicella as a risk factor for iGAS among young children and highlight the association with non-invasive GAS infections in the community as a possible source of transmission.

## Introduction

Group A streptococci (GAS) are gram-positive bacteria (*Streptococcus pyogenes)*, which can be carried asymptomatically or cause non-invasive and invasive disease (iGAS). Non-invasive manifestations of GAS include mild diseases such as impetigo and scarlet fever. Invasive infection is rare, and can lead to severe disease such as meningitis or toxic shock syndrome with high case fatality [1]. There is no vaccine available and public health interventions consist mainly of prescribing antibiotic prophylaxis to household contacts of iGAS patients and monitoring of symptoms among close contacts to allow early treatment [2].

In 2022, an increase of iGAS was observed in the Netherlands, especially among children aged 0-5 years. At that time, three manifestations of iGAS were notifiable in the Netherlands: necrotizing fasciitis, streptococcal toxic shock syndrome (STSS) and puerperal fever or -sepsis [2]. In 2022, 42 cases were reported in the age group 0-5 years; a sevenfold increase compared to pre-COVID years 2016-2019 [3]. Of these 42 cases, 9 deceased. The increase in iGAS was also reported by other European countries such as United Kingdom, France and Ireland [4-9]. The increase could not be fully attributed to a specific *emm-*type, even though there is evidence that the proportion of *emm*1 isolates among children aged 0-5 years increased over the year 2022 [3, 10].

Besides the increase in iGAS infections, a rise in primary care consultations was seen in 2022 for pharyngitis, scarlet fever and varicella zoster [3, 10, 11]. Incidences of viral infections such as influenza and varicella, which are known risk factors for iGAS, were higher than in pre-covid years [10-13]. It is also suggested that infection control measures during the COVID-19 pandemic reduced exposure to GAS and other childhood infections, resulting in a decreased immunity and a higher susceptibility for iGAS [14]. This can be a partial explanation of the surge in iGAS cases during winter season 2022-2023. To identify the risk factors for iGAS among children aged 6 months to 5 years in the post-COVID-19 era, we conducted a prospective case-control study in the Netherlands in early 2023.

## Methods

A prospective case-control study was conducted between 13 February and 30 May 2023 among children aged 6 months to 5 years. A case was defined as a child aged 6 months to 5 years with symptoms of invasive disease and *S. pyogenes* cultured from a normally sterile site or a normally non-sterile site without the presence of another pathogen responsible for the disease, occurring between February and May 2023 in The Netherlands. Parents of all notified cases were invited by the Municipal Health Services (MHS) to participate in the study. If the parents of the case agreed to participate, the online questionnaire was sent by the MHS.

Parents willing to have their children serve as controls were recruited from the general population via social media channels (Facebook, Instagram and LinkedIn) of the National Institute for Public Health and the Environment (RIVM), asking to apply for participation in a survey, which resulted in a pool of potential controls. Controls had to be in the same age range as cases, but without a history of iGAS. In order to take into account the respiratory season, we aimed to have cases and the selected controls filling out the questionnaire around the same moment in time. Whenever a new case was notified, we selected 10 controls from the pool, matched to the case on sex and birthyear, or birth-quarter for children younger than 1 year old. Parents of selected controls were invited via the RIVM by e-mail to fill in the online questionnaire. Controls whose siblings already participated in the study were excluded, as their exposures could not be regarded as independent observations.

The questionnaires’ primary exposure of interest was infectious disease during four weeks prior to disease onset (cases) or during the past 4 weeks (controls). Because we were asking parents, and not clinicians, to answer questions about illness, we added clear descriptions of disease presentations such as scarlet fever, impetigo and varicella. For the analysis, the additional variable any GAS-like illness was created. Any GAS-like illness was defined as scarlet fever or impetigo or pharyngitis. We also created the variable respiratory infection, defined as runny nose or cough or shortness of breath or sore throat. Other exposures of interest were household size, chronic illness, use of medication including antibiotics, going to school or daycare, whether a child visited primary care or emergency care unit, admission to hospital, surgical procedures such as tonsillitis, occurrence of infectious diseases in the child’s social environment and vaccination against COVID-19 or influenza. As a proxy for social economic status highest education level of one of the parents was asked.

Univariable conditional logistic regression was performed to estimate the odds ratios (OR) and 95% confidence intervals (95% CI) of exposures among iGAS cases and controls to identify risk factors for severe disease. Data cleaning and analyses were done in R version 4.2.2 [15].

We verified whether the work complied with the specific conditions as stated in the Dutch Medical Research Law on Human Subjects (WMO). The research does not fulfill one or both of these conditions. Therefore, the study was exempted for further approval by an ethical research committee. A Data Privacy Impact Assessment (DPIA) was conducted to adhere to the data protection legislation. A digital informed consent of parents of cases and controls was requested before the online questionnaire could be opened.

## Results

Of 56 cases notified during the study period, 18 completed the questionnaire (response rate 32%). The control pool existed of 1132 potential controls, of which 578 were matched to a case and therefore invited to fill in the questionnaire. Eighteen additional controls received an invitation erroneously, because of an error in the software used to invite the controls. 351 controls filled in the questionnaire (response rate 61%), with a median of 6 controls per case (interquartile range 4-7). For 248 responding controls, the matched case did not respond, therefore their data could not be included in the primary analysis. An overview of the recruiting process is shown in Figure 1.

**Figure 1.**
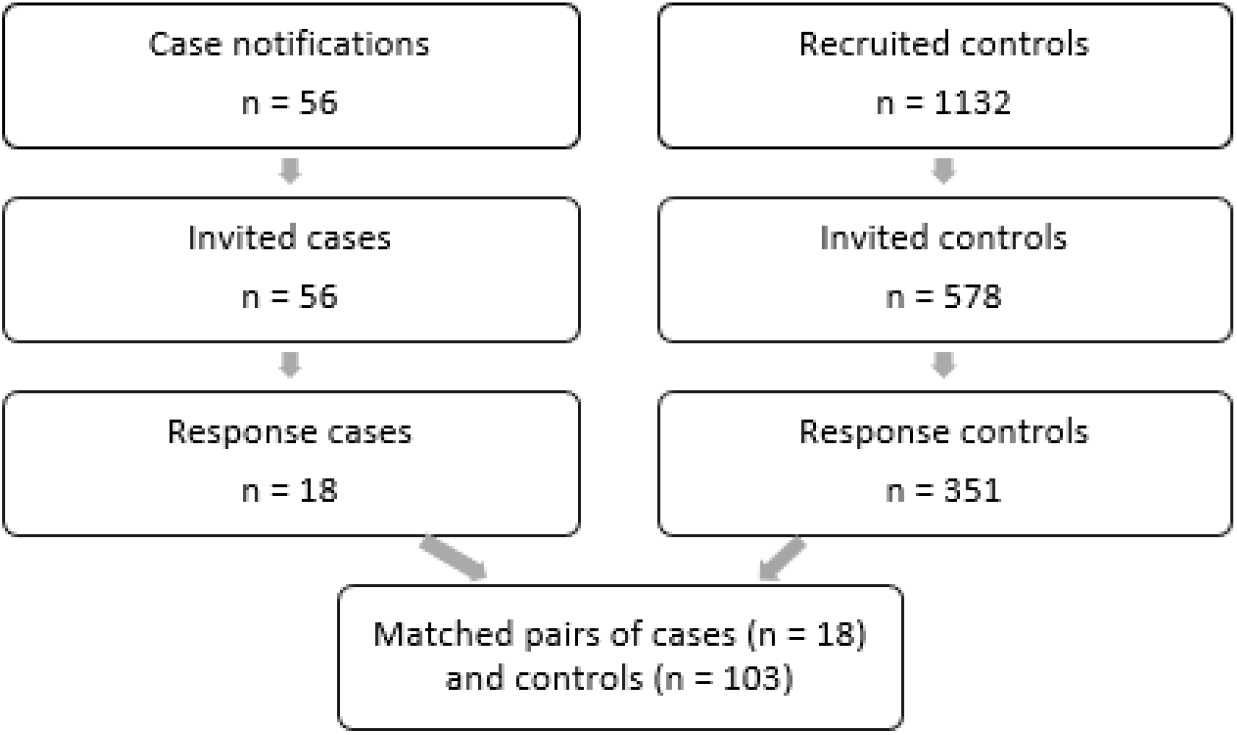
Process from recruitment (February-May 2023) of cases and controls to final dataset for analysis.

For the primary analysis, a total of 18 cases and 103 controls were included. Because of the matching criteria sex and birthyear, the proportions in both groups were similar, see Table 1. Among the cases 10 (56%) were male and 8 (44%) female. Among the controls 55 (53%) were male and 48 (47%) female.

**Table 1.** Matching characteristics of cases and controls.

| Characteristics |  | Cases (N=18) |  | Controls (N=103) |  |
| --- | --- | --- | --- | --- | --- |
|  |  | n | % | n | % |
| Sex | Male | 10 | 56.0 | 55 | 53.0 |
| Year of birth | 2017 | 1 | 5.6 | 7 | 6.8 |
|  | 2018 | 1 | 5.6 | 7 | 6.8 |
|  | 2019 | 3 | 17.0 | 15 | 15.0 |
|  | 2020 | 1 | 5.6 | 8 | 7.8 |
|  | 2021 | 6 | 33.0 | 34 | 33.0 |
|  | 2022 | 6 | 33.0 | 32 | 31.0 |

A complete overview of the analyzed variables can be found in Table 2. Prior respiratory infection was reported among 14 (78%) of cases and 87 (85%) controls, this difference was not statistically significant. Likewise, no statistically significant difference was found between cases and controls for impetigo, pharyngitis and scarlet fever. However, when combining these syndromes into the variable GAS-like illness we found a statistically significant difference with an odds ratio of 8.5 (95% CI: 1.5- 49.0). Prior varicella zoster infection was reported significantly more among cases (OR: 12.0, 95% CI: 1.1-139.0), as was fever (OR: 7.5, 95% CI: 2.1-28.0). None of the included chronic illnesses were significant risk factors. Other factors such as prior use of antibiotics, household size and attending daycare or school were not significant. Medium or high education level of at least one of the parents was associated with a lower risk of developing iGAS (OR: 0.1, 95% CI: 0.0-0.5).

**Table 2.**
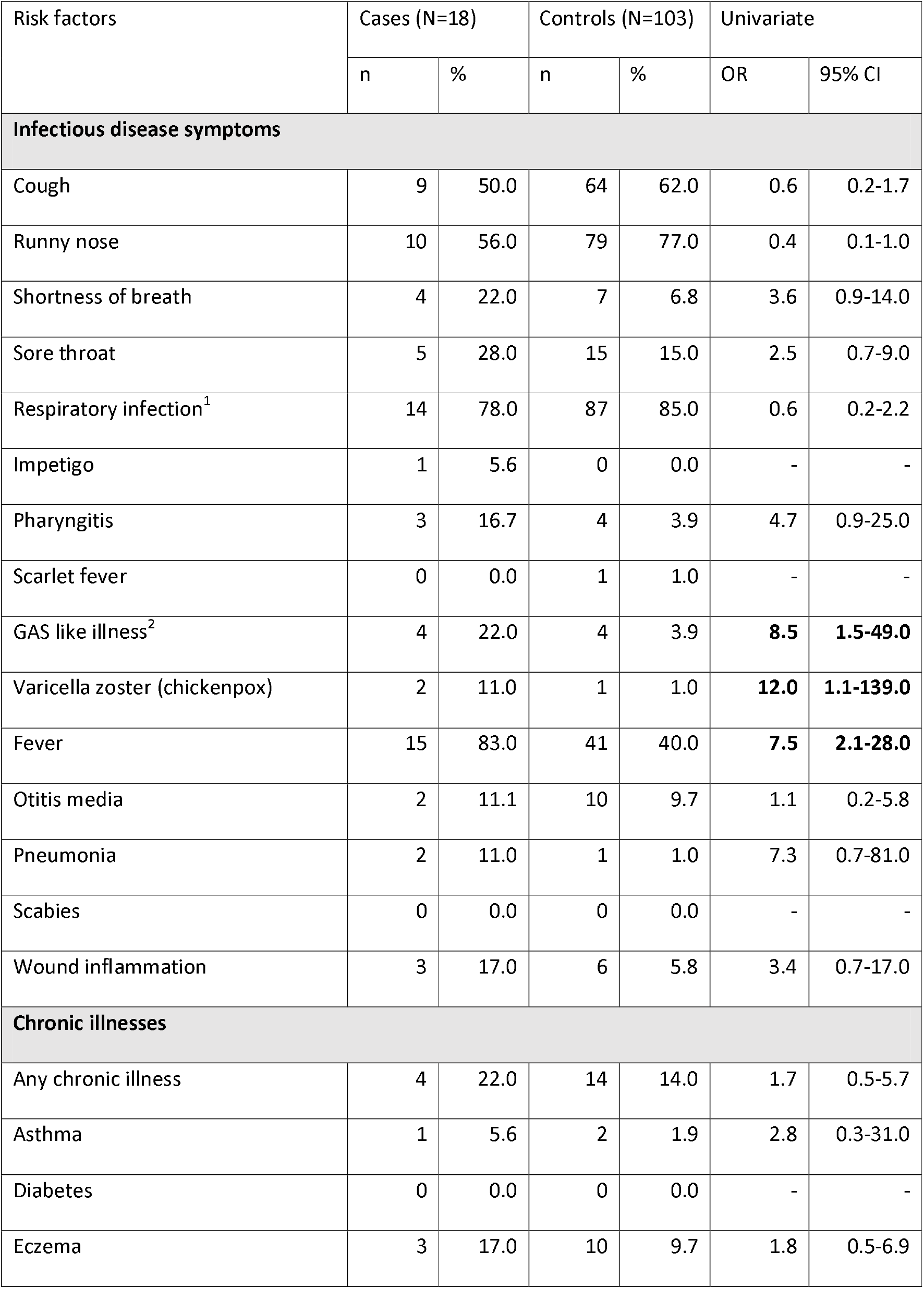
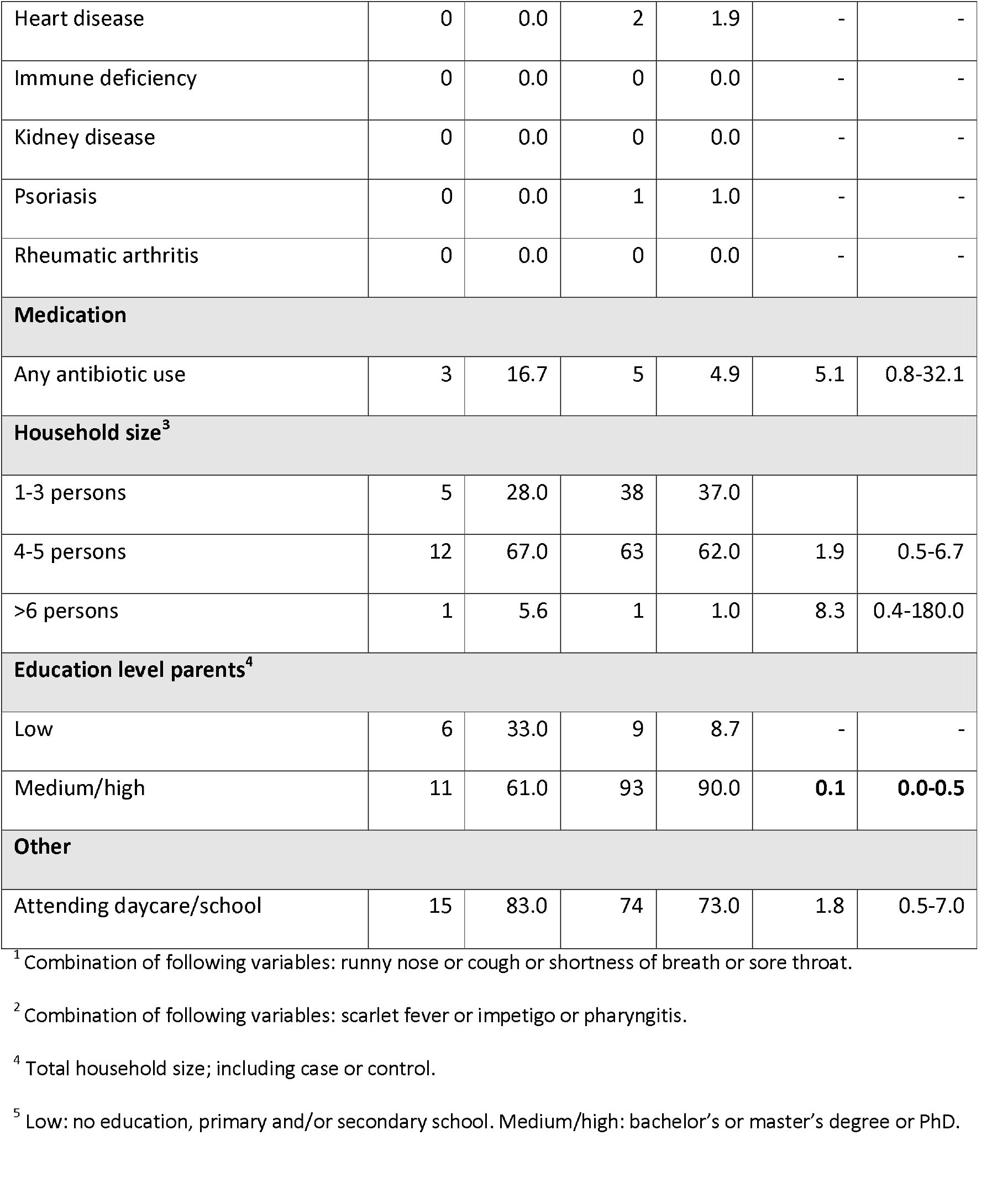
Univariable analysis of potential risk factors for developing iGAS, four weeks prior to disease onset, cases (N = 18) compared to controls (N = 103), Netherlands, February-May 2023.

We also investigated if various infectious disease symptoms were present in the social environment (household, family, friends, daycare or school), of which the GAS-like illnesses had the most significant findings, see Table 3. Three (17%) cases and 1 (1.2%) control reported scarlet fever in their social environment (OR: 24.0, 95% CI: 2.2-257.0). Any GAS like illness was reported in the social environment of 8 (44%) cases and 15 (15%) controls (OR: 7.1, 95% CI: 1.8-29.0). A complete overview of the infectious disease symptoms in the social environment can be found in Supplementary Table 4.

**Table 3.**
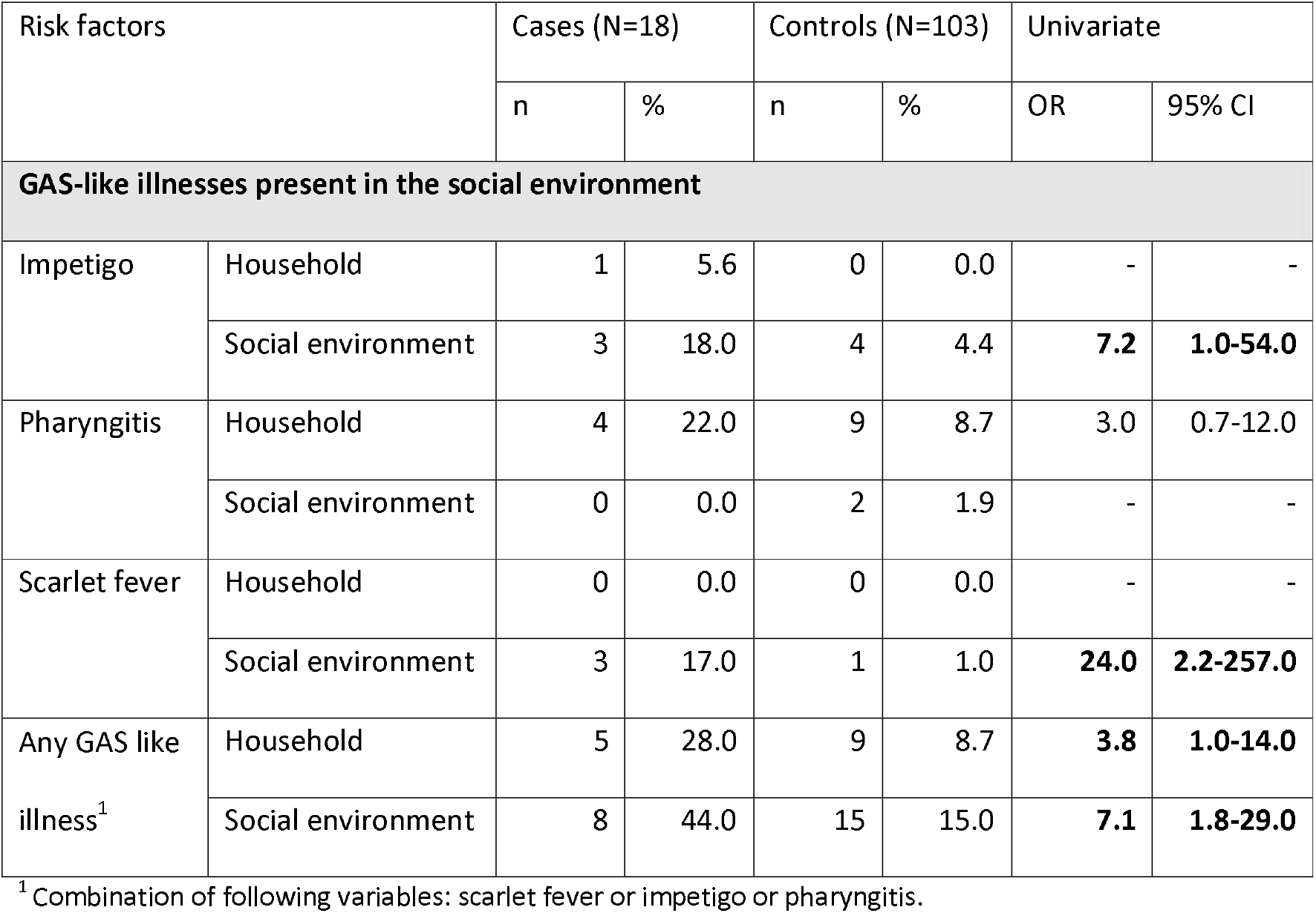
Univariable analysis of GAS-like illnesses in the social environment, four weeks prior to disease onset, cases (N = 18) compared to controls (N = 103), Netherlands, February-May 2023.

| Risk factors |  | Cases (N=18) |  | Controls (N=103) |  | Univariate |  |
| --- | --- | --- | --- | --- | --- | --- | --- |
|  |  | n | % | n | % | OR | 95% CI |
| **GAS-like illnesses present in the social environment** |  |  |  |  |  |  |  |
| Impetigo | Household | 1 | 5.6 | 0 | 0.0 | - | - |
|  | Social environment | 3 | 18.0 | 4 | 4.4 | **7.2** | **1.0-54.0** |
| Pharyngitis | Household | 4 | 22.0 | 9 | 8.7 | 3.0 | 0.7-12.0 |
|  | Social environment | 0 | 0.0 | 2 | 1.9 | - | - |
| Scarlet fever | Household | 0 | 0.0 | 0 | 0.0 | - | - |
|  | Social environment | 3 | 17.0 | 1 | 1.0 | **24.0** | **2.2-257.0** |
| Any GAS like illness<sup>1</sup> | Household | 5 | 28.0 | 9 | 8.7 | **3.8** | **1.0-14.0** |
|  | Social environment | 8 | 44.0 | 15 | 15.0 | **7.1** | **1.8-29.0** |

<sup>1</sup> Combination of following variables: scarlet fever or impetigo or pharyngitis.

### Secondary analysis – varicella zoster

A question about varicella zoster in the 4 weeks prior to disease onset was part of the standard iGAS notification questionnaire. Therefore, this information was available also for cases who did not respond to the questionnaire. We performed a secondary analysis using the data from the notification questionnaire compared to the matched controls. For 47/56 cases, the notification questionnaire contained data about prior varicella zoster infection. A total of 256 control responses were matched to these 47 cases. Prior varicella zoster infection was reported in 6 (13%) cases (2 necrotizing fasciitis, 4 skin and soft tissue infections) and 4 (1.4%) controls (OR: 11.7, 95% CI 3.9-47).

## Discussion

In this study we aimed to identify risk factors for iGAS infection among young children in early 2023. Our findings are in line with previous studies confirming the association between varicella zoster and iGAS and confirm the association with presence of non-invasive GAS infections in the social environment. This latter association was strongest for scarlet fever.

Varicella zoster is known as risk factor for iGAS, mostly associated with GAS skin and soft tissue infection [11]. In early 2022, with re-opening of society after multiple lockdowns, varicella zoster infections surged after two years of low incidence and a likely build-up of susceptible individuals, in the absence of varicella vaccination in the Netherlands. This provided a likely explanation for the peak in iGAS among young children observed in the Netherlands in early 2022 [3]. After the varicella zoster peak in 2022, 2023 showed a remarkably low varicella zoster incidence [16]. Still, in both our primary and our secondary analysis, we found a strong association between varicella zoster and iGAS infection. Contact with persons with scarlet fever, impetigo or pharyngitis has previously been described as a risk factor for developing iGAS [17, 18]. We found the strongest association between iGAS infection and scarlet fever, the only syndrome in the questionnaire that is specifically caused by GAS infection. Our findings are consistent with the notion that these non-invasive GAS infections can be important sources of transmission [17, 18]. Respiratory viral infections, specifically influenza, have been described previously as a risk factor for iGAS as well [12, 13]. However, in our study there was no statistically significant difference in symptoms of respiratory infection between cases and controls. The interpretation of our finding of fever as a risk factor is difficult, because this may have been an early symptom of the iGAS rather than a prior infection by another pathogen.

A strength of this study is the quick approach of parents of cases via the MHS to fill in the questionnaire which likely minimized recall bias. Also the call for participation of controls via social media was an efficient method to recruit enough controls to conduct the study. Our prospective design, with recruitment of controls at the study onset but sending the questionnaire only when a matched case was notified, allowed us to study various seasonal risk factors without confounding by calendar time. This method can be used to quickly deploy case-control studies in future outbreaks where time-varying risk factors are likely to play a role.

One of the main limitations of this study is the low number of included cases. iGAS infection among young children remains a rare disease, and our study commenced almost a year after the start of the iGAS increase in the Netherlands. Also, the majority of the approached parents of cases chose not to participate. Parents of cases were approached during a stressful period when their child was hospitalized, and in some cases deceased. Understandably, many of these parents did not have the mind space for participation. This may also have introduced some selection bias towards cases with less severe disease, as we know that at least five cases deceased but none of them were part of this case-control study. We also suspect some selection bias in the recruitment and selection of controls. We recruited participants through RIVM social media channels to sign up for the study; this might have reached a specific highly educated, and health- and research-minded population, which is not representative for the total population in the Netherlands. This may explain the significantly higher education level among parents of controls.

Due to the small numbers we could not conduct a thorough multivariable analysis and correct for possible confounders such as household size, education level and co-morbidities. Another limitation is that the risk factors in this study were only identified by anamnesis of the parents, and not proven by any microbiological analysis.

## Conclusion

This study showed that the risk of iGAS in children 6 months to 5 years is associated with prior varicella zoster infection and exposure to GAS like illnesses in the social environment, in particular scarlet fever. General practitioners should be aware of these pediatric iGAS risk factors, especially in settings of high varicella zoster and/or scarlet fever circulation. The results of this study are relevant for informing policies on varicella zoster vaccination and distribution of post-exposure prophylaxis among contacts of iGAS cases to prevent invasive disease, especially in outbreak settings where multiple pathogens are co-circulating and increasing the risk of developing iGAS.

## Data Availability

All data produced in the present study are available upon reasonable request to the authors, in aggregated form to ensure participant privacy.

## Acknowledgements

The authors would like to thank all participating parents for contributing to this study, the Municipal Health Services in the Netherlands for their support in the data collection, the OSIRIS support team and the Infectieradar-team for using their platform to distribute questionnaires.

