## Supplementary Table 4 for "Risk factors for invasive Group A Streptococcal infection in children aged 6 months to 5 years, a case-control study, the Netherlands, February-May 2023"

**Supplementary material**

Supplementary table S4. Univariable analysis of symptoms of infectious diseases present in the social environment, four weeks prior to disease onset, cases (N = 18) compared to controls (N = 103), Netherlands, February-May 2023.

| Risk factors | | Cases (N=18) | | Controls (N=103) | | Univariate | |
| --- | --- | --- | --- | --- | --- | --- | --- |
|  |  | n | % | n | % | OR | 95% CI |
| Infectious disease present in the social environment | | | | | | | |
| Cough | Household | 4 | 23.5 | 55 | 53.4 | 0.3 | 0.1-0.9 |
|  | Social environment | 4 | 23.5 | 71 | 70.3 | 0.1 | 0.0-0.4 |
| Runny nose | Household | 7 | 41.2 | 71 | 68.9 | 0.4 | 0.1-1.0 |
|  | Social environment | 9 | 52.9 | 81 | 78.6 | 0.3 | 0.1-1.4 |
| Shortness of breath | Household | 1 | 5.9 | 9 | 8.7 | 0.7 | 0.1-5.9 |
|  | Social environment | 1 | 5.9 | 17 | 18.3 | 0.4 | 0.1-3.6 |
| Sore throat | Household | 7 | 41.2 | 39 | 37.9 | 1.2 | 0.4-3.3 |
|  | Social environment | 7 | 38.9 | 52 | 52.0 | 0.9 | 0.2-3.2 |
| Respiratory infection | Household | 11 | 61.1 | 76 | 73.8 | 0.6 | 0.2-1.6 |
|  | Social environment | 12 | 66.7 | 86 | 83.5 | 0.5 | 0.1-2.4 |
| Impetigo | Household | 1 | 5.6 | 0 | 0.0 | - | - |
|  | Social environment | 3 | 18.0 | 4 | 4.4 | 7.2 | 1.0-54.0 |
| Pharyngitis | Household | 4 | 22.0 | 9 | 8.7 | 3.0 | 0.7-12.0 |
|  | Social environment | 0 | 0.0 | 2 | 1.9 | - | - |
| Scarlet fever | Household | 0 | 0.0 | 0 | 0.0 | - | - |
|  | Social environment | 3 | 17.0 | 1 | 1.0 | 24.0 | 2.2-257.0 |
| GAS like illness | Household | 5 | 28.0 | 9 | 8.7 | 3.8 | 1.0-14.0 |
|  | Social environment | 8 | 44.0 | 15 | 15.0 | 7.1 | 1.8-29.0 |
| Varicella zoster | Household | 2 | 11.8 | 0 | 0.0 | - | - |
|  | Social environment | 3 | 16.7 | 6 | 6.7 | 5.0 | 0.9-27.4 |
| Fever | Household | 5 | 29.4 | 25 | 24.3 | 1.3 | 0.4-3.8 |
|  | Social environment | 5 | 27.8 | 43 | 43.9 | 0.6 | 0.2-2.3 |
| Otitis media | Household | 3 | 16.7 | 7 | 6.8 | 2.7 | 0.6-11.6 |
|  | Social environment | 3 | 17.6 | 14 | 15.9 | 1.3 | 0.3-6.5 |
| Pneumonia | Household | 0 | 0.0 | 1 | 1.0 | - | - |
|  | Social environment | 0 | 0.0 | 2 | 2.3 | - | - |
| Scabies | Household | 0 | 0.0 | 0 | 0.0 | - | - |
|  | Social environment | 0 | 0.0 | 0 | 0.0 | - | - |
| Wound inflammation | Household | 1 | 5.9 | 2 | 1.9 | 2.7 | 0.2-30.6 |
|  | Social environment | 1 | 5.9 | 4 | 4.5 | 1.5 | 0.1-17.3 |

^1^ Combination of following variables: runny nose or cough or shortness of breath or sore throat.
^2^ Combination of following variables: scarlet fever or impetigo or pharyngitis.
